# Priority age targets for COVID-19 vaccination in Ethiopia under limited vaccine supply

**DOI:** 10.1101/2022.07.28.22278142

**Authors:** Margherita Galli, Agnese Zardini, Worku Nigussa Gamshie, Stefano Santini, Ademe Tsegaye, Filippo Trentini, Valentina Marziano, Giorgio Guzzetta, Mattia Manica, Valeria d’Andrea, Giovanni Putoto, Fabio Manenti, Marco Ajelli, Piero Poletti, Stefano Merler

## Abstract

**Background:** The worldwide inequitable access to vaccination claims for a re-assessment of policies that could minimize the COVID-19 burden in low-income countries. An illustrative example is what occurred in Ethiopia, where nine months after the launch of the national vaccination program in March 2021, only 3% of the population received two doses of COVID-19 vaccine. In the meantime, a new wave of cases caused by the emergence of Delta variant of SARS-CoV-2 was observed between July and November 2021.

**Methods:** We used a SARS-CoV-2 transmission model to estimate the level of immunity accrued before the launch of vaccination in the Southwest Shewa Zone (SWSZ) and to evaluate the impact of alternative age priority vaccination targets in a context of limited vaccine supply. The model was informed with available epidemiological evidence and detailed contact data collected across different socio-demographic settings.

**Results:** We found that, during the first year of the pandemic, 46.1-58.7% of SARS-CoV-2 infections and 24.9-48% of critical cases occurred in SWSZ were likely associated with infectors under 30 years of age. During the Delta wave, the contribution of this age group in causing critical cases was estimated to increase to 66.7-70.6%. However, our findings suggest that, when considering the vaccine product available at the time (ChAdOx1 nCoV-19; 65% efficacy against infection after 2 doses), prioritizing the elderly for vaccination remained the best strategy to minimize the disease burden caused by Delta, irrespectively to the number of available doses. Vaccination of all individuals aged 50□years or older would have averted 40 (95%CI: 18-60), 90 (95%CI: 61-111), and 62 (95%CI: 21-108) critical cases per 100,000 residents in urban, rural, and remote areas, respectively. Vaccination of all individuals aged 30□years or more would have averted an average of 86-152 critical cases per 100,000 individuals, depending on the setting considered.

**Conclusions:** Despite infections among children and young adults likely caused 70% of critical cases during the Delta wave in SWSZ, most vulnerable ages should remain a key priority target for vaccination against COVID-19.

## Background

Two years into the pandemic, the reported burden of the coronavirus disease 2019 (COVID-19) has been relatively low throughout Africa as compared to high-income countries [1,2]. In Africa, approximately 40% of people are aged less than 15 years, compared to a global mean of 25% [3], and severe outcomes of COVID-19 are strongly associated with age [4-6]. However, the impact of COVID-19 in low-income countries may have been vastly underestimated due to lacking testing capacity [7-9]. For instance, a recent post-mortem study in Zambia revealed that, contrary to expectations, deaths possibly ascribable to COVID-19 were common among patients of a referral hospital, with about 20% deceased individuals resulting infected with SARS-CoV-2 compared to less than 10% tested before death [10].

The identification of appropriate strategies to minimize COVID-19 burden in sub-Saharan settings remains an open challenge. Unprecedented social distancing measures have been applied worldwide to mitigate the COVID-19 pandemic [11-15]. However, the implementation of drastic restrictions for long time periods would have disproportionate effects on the already vulnerable economies of low-income countries [12,13,15]. Mass immunization programs still represent the main public health strategy to reduce COVID-19 burden. While high-income countries have rapidly progressed in the deployment of multiple vaccine doses, at the end of 2021, only 15% of the total African population was vaccinated with at least one dose [1].

Ethiopia represents an illustrative case study for the limited access to vaccination experienced by sub-Saharan countries during 2021. In this country, the national vaccination campaign was launched on March 13, 2021 [16], with a focus on high-risk categories (i.e., elderly, patients with chronic diseases, and health care workers). On November 16, 2021, the vaccination campaign was expanded to all individuals aged 12 years or more. Nonetheless, the vaccine uptake of Ethiopia has remained negligible for a long period of time, with only 3.1% of the citizens being vaccinated with two doses after nine months from the start of vaccination [1,17].

In this study, we assess the potential impact of different vaccination policies in reducing the burden caused by the Delta variant of SARS-CoV-2 across different socio-demographic settings of the Southwest Shewa Zone (SWSZ) of Ethiopia in the context of limited vaccine supply. Alternative priority targets for vaccination are evaluated by considering different scenarios regarding the available number of vaccine doses and by taking into account the immunity acquired by natural infection before the launch of the national vaccination campaign. To do this, we develop and simulate a transmission model for SARS-CoV-2 informed with data on age-specific mixing patterns recently collected across different areas of the SWSZ, characterized by heterogeneous population density, age structure, and access to primary care [11]. The effect of different immunization strategies is evaluated in terms of the number of infections and critical cases that could have been averted after the rollout of vaccination based on ChAdOx1 nCoV-19, representing the vaccine predominately adopted during 2021 in Ethiopia. Obtained results could be instrumental to identify the optimal strategies for the deployment of vaccines in socio-economic contexts characterized by an initial limited vaccine supply.

## Methods

The SARS-CoV-2 transmission dynamics is simulated by using a deterministic age-structured SIR model. Susceptibility to SARS-CoV-2 infection is assumed to vary with age according to estimates made available by Hu et al. [18]. Specifically, taking the age group of 20-59 years as the reference, the relative susceptibility for individuals aged 0-19 years is set at 0.59 (95%CI: 0.35-0.92) and at 1.75 (95%CI: 1.07-2.81) for the individuals aged 60 years or more. An average generation time of 6.6 days and homogenous infectiousness across different ages are assumed [19,20].

The developed model keeps track of the contribution of infectors of different ages in causing secondary infections and critical cases across different socio-demographic contexts. Critical disease cases are defined as positive individuals who would either require intensive care or result in a fatal outcome. Age-specific risks of developing critical disease after SARS-CoV-2 infection are considered [4]. The adopted approach leverages on age-specific contact matrices recently estimated for rural villages, dispersed subsistence farming settlements, and urban neighborhoods of the SWSZ of the Oromia Region, Ethiopia [11]. The model is run separately for each geographical context, assuming a constant population size over time, and accounting for the age structure characterizing the settings under study (urban, rural, and remote) [11].

The contribution of different ages in causing secondary infections and critical cases is explored by considering two pandemic phases. As for the first phase, lasting until the launch of the national vaccination program in March 2021, we consider the emergence of SARS-CoV-2 in a fully naïve population of individuals and analyze the epidemic dynamics under the dominance of the ancestral strain of SARS-CoV-2 and in the absence of vaccination. A school closure mandate is also assumed for the entire period as this represented a persistent restriction adopted by the government to counter the spread of COVID-19 during the first pandemic wave [11,21]. The spread of infection is simulated by considering an initial reproduction number of 1.62 (95%CI: 1.55–1.70), as estimated from the exponential growth of cases reported in Ethiopia from May to mid-June 2020 [11]. This corresponds to assuming for the ancestral strain a basic reproduction number (R_0_) around 3, which is in line with estimates available from other countries [22]. The transmission dynamics during this pandemic phase is simulated until a given setting specific proportion of the population gets infected. Such proportion is defined according to the levels of serological prevalence estimated for March 2021 in the Jimma Zone of Ethiopia: 31% in rural and remote sites and 45% in urban areas [23]. Different seroprevalence values are considered for sensitivity analysis to account for the uncertainty surrounding the circulation of the infection before March 2021 and the potential waning of naturally acquired immunity. The model ability in capturing the observed epidemiological patterns is assessed by comparing the age distribution of the cumulative number of simulated infections with the one associated with SARS-CoV-2 positive individuals ascertained with real-time reverse transcription polymerase chain reaction (RT-PCR) between March and September 2020 in the Oromia Region [24].

The second pandemic phase that we consider mirrors the SARS-CoV-2 transmission dynamics after the launch of the national vaccination program in March 2021, when students were regularly receiving in-person education [21]. Model estimates of the natural immunity acquired by different age groups during the first pandemic phase are used to initialize the immunological status of the population in this second phase. To account for the replacement of the ancestral lineages by the Delta variant of SARS-CoV-2 likely occurred in mid 2021 [25], we assume a transmission rate mirroring a R_0_=6 [26]; alternative values of R_0_ are explored for sensitivity analysis.

The impact of different vaccination strategies on the burden of COVID-19 is assessed in terms of the potential attack rate of infection and the cumulative incidence of critical cases expected after March 2021, in the absence of restrictions on the individuals’ contacts. The comparison of alternative vaccination priority groups is carried out by assuming that the considered vaccination target is achieved before the upsurge of cases caused by the emergence of the Delta variant, neglecting the transient dynamic characterizing the rollout of the vaccination.

Four illustrative scenarios are analyzed. First, we consider a scenario where the number of administered vaccines is negligible, and we evaluate the impact of pre-existing immunity levels in shaping the contribution of different ages to the disease spread. Given the low vaccine uptake recorded in Ethiopia, this scenario may reflect what might have occurred in the months following the launch of vaccination because of Delta expansion in the population. Second, we assume that a limited number of doses is available, and we compare the vaccination program targeting 100% of individuals aged 50□years or older, representing the initial age priority target defined by the Ethiopian vaccination program [27], with an alternative scenario where the corresponding number of vaccine doses is offered to all ages eligible for vaccination (≥10 years of age). Third, we assume that all individuals aged 50 years or more are fully vaccinated and we project the potential impact of expanding vaccination to other age groups. In this case, we compare the impact of administering the vaccine only to individuals aged 30-49 years with an alternative scenario where the corresponding number of doses is uniformly distributed to all eligible ages (10-49 years). Finally, to provide a comprehensive view of the potential benefits of vaccination, we consider different combinations of coverage levels attained among subjects aged 50 years or more and individuals aged between 10 and 49 years, irrespectively of the number of doses and logistic efforts required to achieve the considered targets.

In the model, vaccinated individuals are assumed to receive two doses of vaccine which significantly reduce their risk of infection and of developing severe outcomes [28-34]. Since ChAdOx1 nCoV-19 was the dominant vaccine employed in Ethiopia during 2021 [35], the vaccine efficacy against infection and critical diseases is set at 65% and 71.5%, respectively [30-34,36]. In a sensitivity analysis, different values for the vaccine efficacy are considered to reflect the use of alternative vaccine products, the administration of only one dose of the vaccine, and a lower vaccine effectiveness against the Delta variant caused by the progressive waning of vaccine-induced protection [37]. The SARS-CoV-2 infectiousness of breakthrough infections (i.e., infections occurring among vaccinee) is assumed to be reduced by 50% [28].

Epidemiological transitions are modeled by the following system of ordinary differential equations:

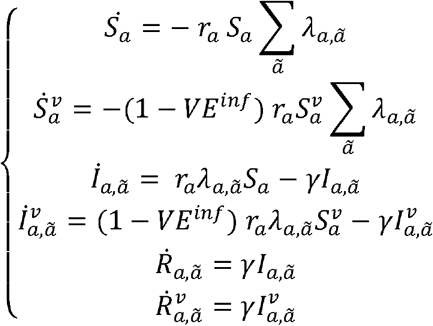

where *a* defines the age of the individuals, *S*_*a*_ represents susceptible individuals of age *a* who have never been vaccinated, 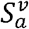 represents vaccinated individuals of age *a* who experienced a reduced force of infection, *VE*^*inf*^ is the vaccine efficacy against the infection, *l*_*a ã*_ and 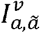 represent the unvaccinated and vaccinated individuals of age *a* infected by subjects of age *ã, R*_*a,ã*_ and 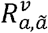 represent the corresponding number of individuals who recovered from these two classes, *r*_*a*_ is the relative susceptibility in the age class 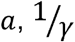 is the average duration of the infectivity period. Finally, *λ*_*a,ã*_ represents the contribution of age *ã* to the force of infection experienced by susceptible individuals of age *a*, which is defined as follows:

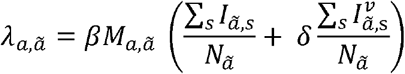

where *M*_*a,ã*_ represents the average number of daily contacts that an individual of age class *a* has with persons of age group *ã, β* is a scaling factor shaping the SARS-CoV-2 transmission rate, *N*_*ã*_ is the total population in the age class *ã*, and *δ* is the relative infectiousness of vaccinated cases, hereafter assumed to be 0.5.

## Results

### SARS-CoV-2 transmission in the pre-vaccination period

The age distribution of the infections estimated with the model under the assumption of a fully susceptible population and by considering the school closure mandate well compares with the one associated with SARS-CoV-2 infections ascertained via PCR in the Oromia Region between March and September 2020 [24] (Figure 1A). Results obtained on the spread of SARS-CoV-2 before the start of COVID-19 vaccination (March 2021) suggest a marked variability across the different geographical contexts in the expected proportion of individuals who acquired natural immunity over 50 years of age: from 47.6% (95%CI: 37.5-59.9%) in rural areas to 64.6% (95%CI: 48.4-78.9%) in the remote settlements (Figure 1B). Our estimates of serological profiles also show a relatively higher immunity among individuals under 50 years of age in urban neighborhoods compared to other settings.

**Figure 1.**
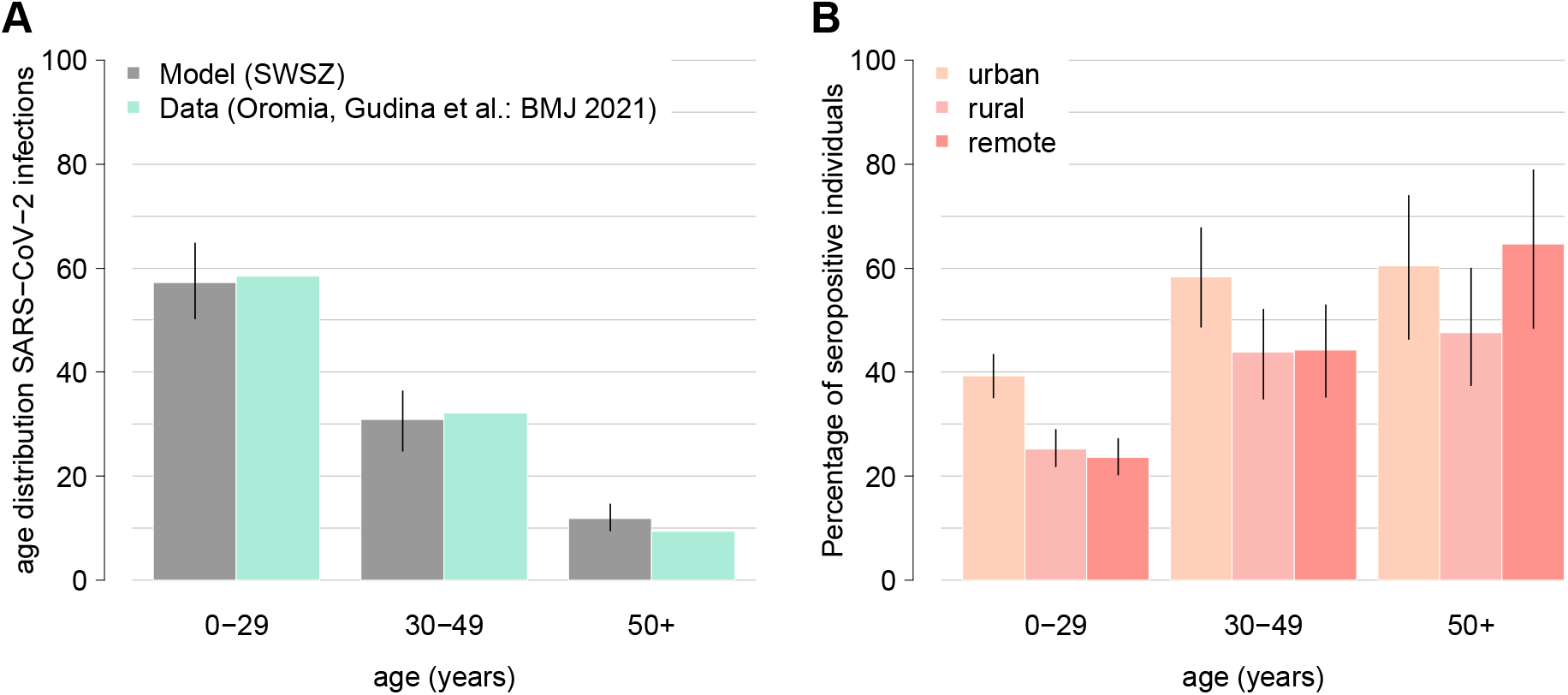
**A)** Comparison between the age distribution of all confirmed cases reported between March and September 2020 in the Oromia Region [24] and the age distribution of the cumulative infections as obtained with a model mimicking the school closure and the achievement of immunity levels estimated for the Jimma Zone in March 2021 [23]. Aggregated model estimates for the entire SWSZ are obtained by considering the proportion of population living in remote settlements, rural villages, and in urban neighborhoods of the SWSZ, their age structure, and the age-specific infection attack rate expected across the different social contexts before March 2021 [11]. **B)** Model estimates of the age-specific percentage of the population immune to SARS-CoV-2 after natural infection at the beginning of the vaccination campaign (March 2021) in urban, rural, and remote areas of the SWSZ. Colored bars represent average estimates; solid lines represent the 95% CI of model estimates.

According to our simulations, the highest fraction of SARS-CoV-2 infections during the first pandemic year was caused by infectors aged less than 30 years: 46.1-58.7% across all the considered socio-economic contexts (see Figure 2B). Infectors younger than 30 years of age might have been responsible for 24.9-48% of critical cases. However, a non-negligible fraction of transmission was found to be assortative, i.e., characterized by a similar age between the infectors and their secondary cases. Specifically, we estimate that, in the remote settlements, 48.7% of infections over 60 years of age might have occurred because of social interactions occurred within this age group. In this setting, individuals aged 50 years or more might have caused half of all critical cases (50.9% in all ages vs 15.9-18.9% in the urban and rural areas, see Figure 2B). This may be explained by the older population structure characterizing less urbanized populations, and the higher number of community contacts reported by the elderly with individuals of similar age (see Figures S1 and S2).

**Figure 2.**
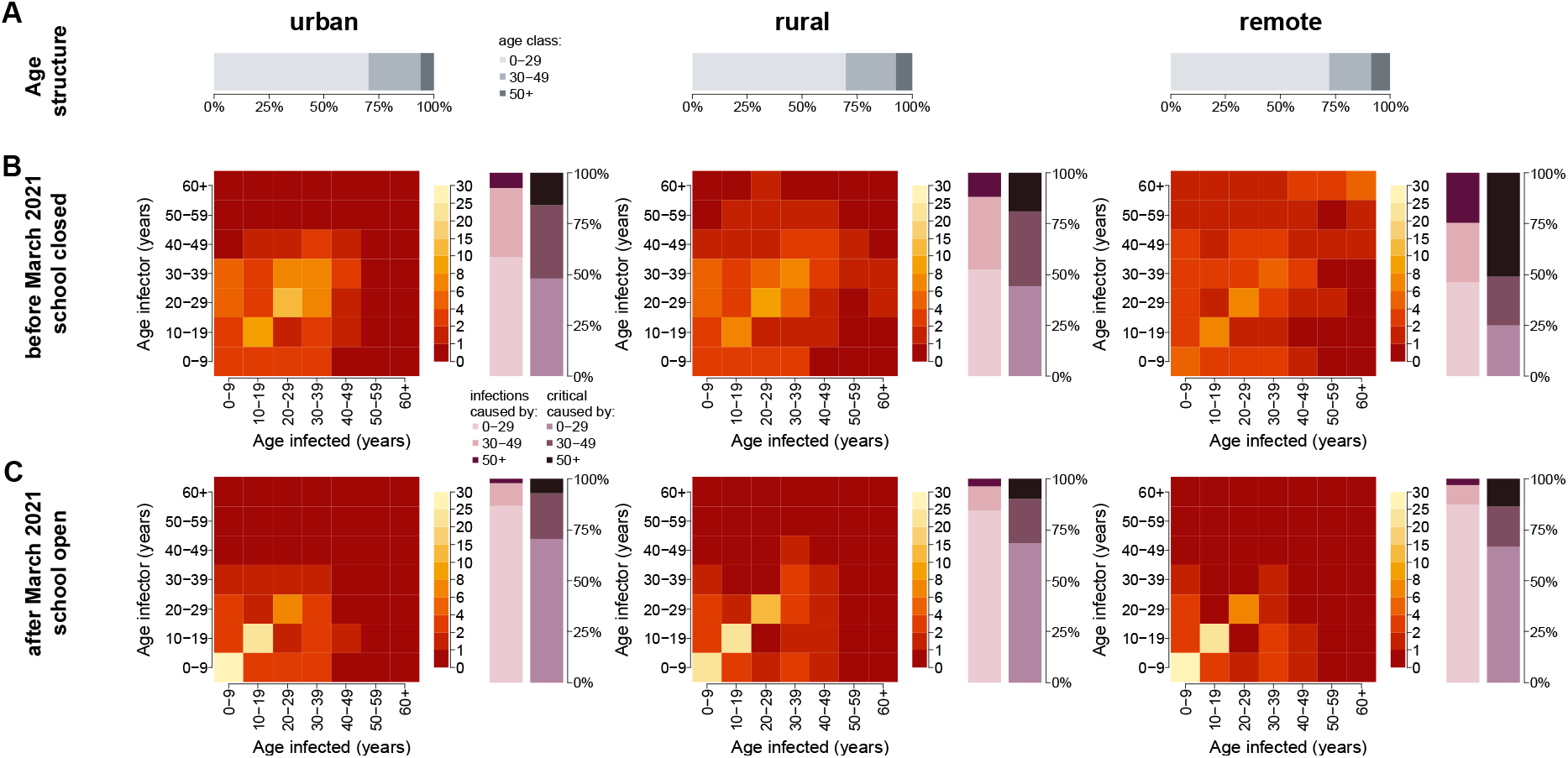
**A)** Age distribution of the population residing in urban neighborhoods, rural villages, and remote settlements. **B-C)** Percentage of SARS-CoV-2 infections caused by contacts between susceptible individuals in the age group *a* (x axis) and infected individuals in the age group *ã* (y axis), as estimated by the model before and after March 2021; bar plots represent the overall proportion of infections and critical cases caused by infectors aged 0-29, 30-49, 50+ years.

### SARS-CoV-2 transmission at vaccination launch

To mimic the COVID-19 epidemiology during the emergence of the Delta variant, we simulate the SARS-CoV-2 transmission under the assumption that the vaccine uptake achieved in the entire population was negligible. However, pre-existing immunity levels as estimated for March 2021 are considered and an increased viral transmissibility is assumed to reflect the transmission advantage of the Delta variant compared to pre-circulating strains [26]. Our results suggest that the natural immunity acquired in the first pandemic phase and the reopening of teaching activities would have reshaped the contribution of different ages in the spread of COVID-19 (see Figure 2C). Specifically, we find that, after March 2021, the contribution of individuals under 30 years of age in causing new infections and critical cases might have increased to 84.5-87.3% and 66.7-70.6%, respectively. Accordingly, we estimate a decrease in the contribution of the elderly in generating SARS-CoV-2 secondary infections and critical cases to 2.0-3.5% and 7.2-13.5%, respectively.

Our estimates suggest that, as the fraction of vaccinated individuals has remained negligible until December 2021, the cumulative incidence of critical cases expected during the Delta wave might have reached 134 (95%CI: 91-174), 223 (95%CI: 180-259), 173 (95%CI: 118-234) per 100,000 residents in the urban, rural, and remote settings, respectively.

### Epidemiological outcome considering different vaccine uptake and priority targets

Our findings suggest that, with a limited vaccine supply, the best strategy to reduce the potential burden of critical disease is to prioritize vaccination of older individuals (see Figure 3C). Specifically, we find that the vaccination of 100% of individuals aged 50 years or older has the potential of averting 40 (95%CI: 18-60), 90 (95%CI: 61-111), and 62 (95%CI: 21-108) critical cases per 100,000 residents in urban, rural, and remote areas, respectively. If the same number of vaccine doses would be uniformly administered to individuals aged 10 years or more, the average number of averted critical cases is expected to be in the range of 11-22 per 100,000 residents, depending on the geographical context considered (see Figure 3).

**Figure 3.**
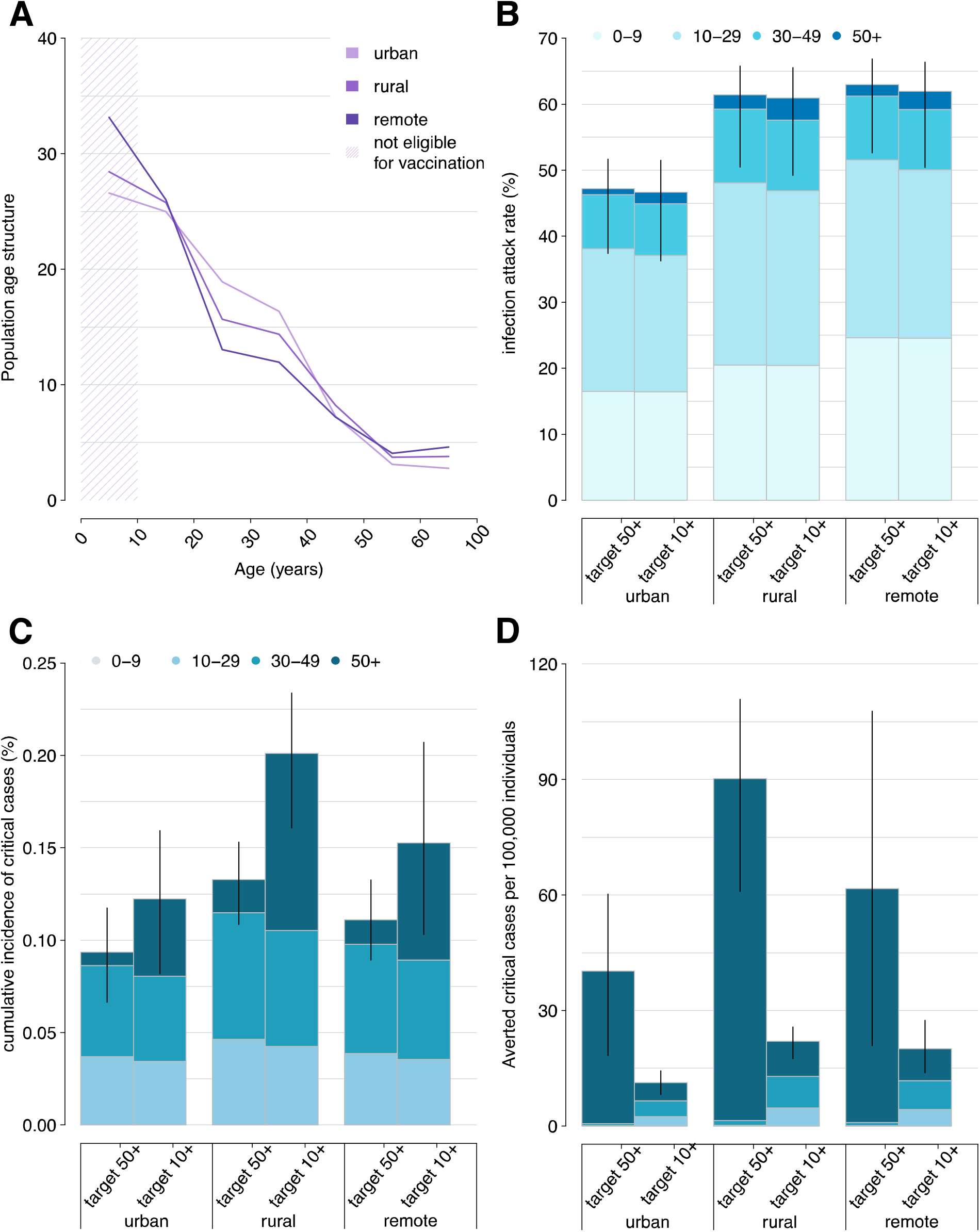
**A**) Population age structure in urban, rural, and remote settings of the SWSZ. The shaded area highlights the age segments of the population who are not yet eligible for COVID-19 vaccination. **B**-**D**) Infection attack rate, cumulative incidence of critical cases, and averted critical cases per 100,000 residents as estimated for different geographical contexts (urban, rural, and remote) under the assumption that either all the individuals aged 50 years or older are vaccinated or that the corresponding number of vaccine doses is uniformly distributed throughout the population over 10 years. Bars represent average estimates, stratified by the age group of infected individuals (0-9, 10-29, 30-49, 50+ years); solid lines represent the 95% CI of model estimates.

As concerns the reduction of the number of infections, the two alternative vaccination strategies result almost equivalent, with differences in the expected attack rates ranging from 0.5% to 1.1% across the three geographical contexts (see Figure 3).

We then explore the scenario where vaccination is expanded to younger age groups after all individuals over 50 years of age are fully vaccinated. We find that the best vaccination policy to further reduce the burden of critical cases remains prioritizing the older segments of the population (i.e., people aged between 30 and 49 years, see Figure 4B). Compared to a scenario with no vaccination, administering the vaccine to all individuals aged 30 years or more would avert 86 (95%CI: 56-113), 152 (95%CI: 120-181), 114 (95%CI: 68-164) critical cases per 100,000 residents in urban, rural, and remote areas, respectively. This policy is estimated to halve the cumulative incidence of critical disease otherwise expected if only individuals older than 50 years get the vaccine (0.05-0.07% vs 0.09-0.13%). Our estimates suggest that the most effective strategy to reduce the infection attack rate is to uniformly distribute the available vaccines among individuals aged 10-49 years. However, the percentage of infections averted under this policy is limited to less than 10% across all considered contexts (see Figure 4A).

**Figure 4.**
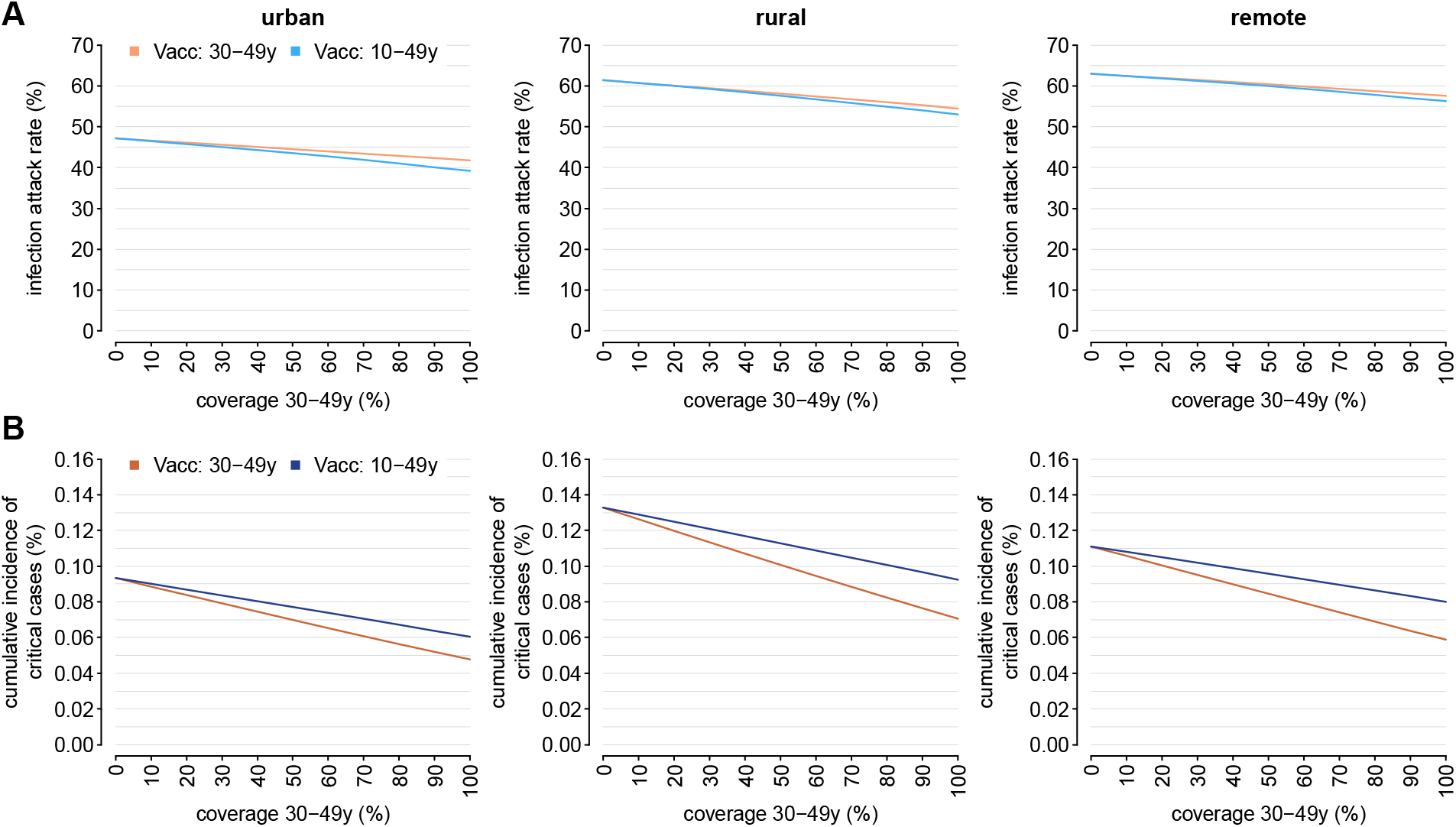
**A**) Estimated infection attack rate in urban, rural, and remote areas, as obtained under the assumption that all individuals above 50 years are vaccinated with two doses and by considering different scenarios for the number of additional doses that would be available. In each panel, two strategies are compared: in the first, a further vaccination effort is simulated to reach a specific coverage level in subjects aged 30-49 years (orange); in the second, the same number of doses is used to uniformly vaccinate individuals aged 10-49 years (blue). Solid lines represent the mean model estimates; shaded areas represent the 95% CI. **B)** As **A)** but for the estimated cumulative incidence of critical cases.

To illustrate the full potential of COVID-19 vaccination, we finally estimate the infection attack rate and the cumulative incidence of critical cases under different combinations of vaccination coverage in the elderly (≥50 years of age) and in individuals aged 10-49 years, irrespectively to possible limits in the vaccine supply and logistic constraints (see Figure 5). Obtained results confirm that the most effective strategy to reduce the number of SARS-CoV-2 infections is the vaccination of younger subjects. However, our estimates suggest that the vaccination of the entire population over 10 years with 2 doses of ChAdOx1 nCoV-19 would have not been enough to decrease the reproduction number below the critical threshold of 1, therefore suggesting that further efforts would have been required to interrupt the SARS-CoV-2 circulation in Ethiopia.

**Figure 5.**
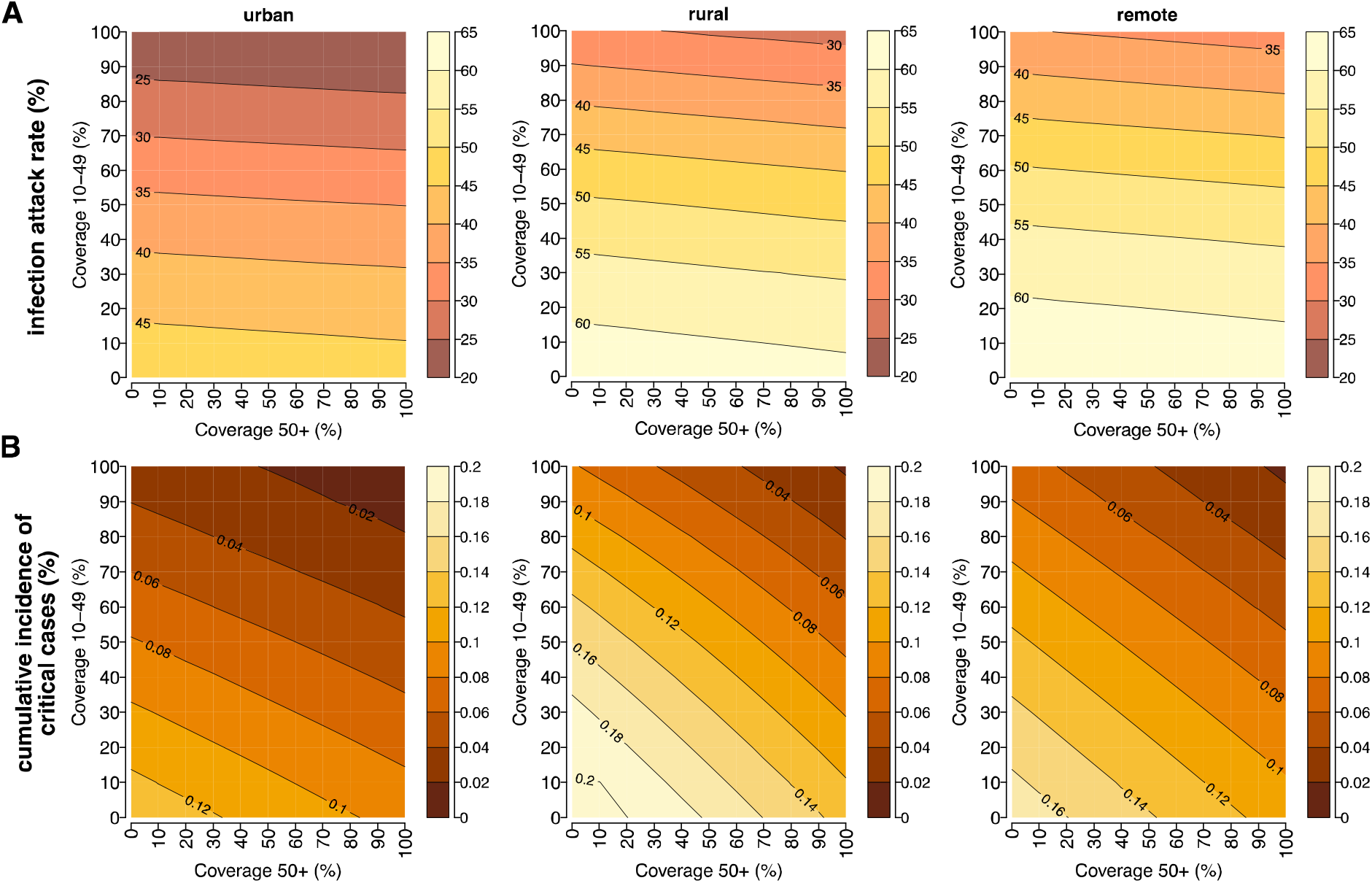
**A)** Infection attack rate as estimated for urban, rural, and remote areas for different combinations of vaccination coverage in individuals aged 50 years or more and in individuals younger than 50 years. **B)** As **A)** but for the estimated cumulative incidence of critical cases (%).

When assuming that all individuals aged 50 years or more are vaccinated, the lowest cumulative incidence of critical cases is estimated to occur in urban neighborhoods, where 93 (95%CI: 66-118) subjects per 100,000 residents are estimated to be exposed to COVID-19 critical disease (see Figure 5B). To reduce the number of critical cases in rural areas under such an incidence level, the strategy minimizing the number of administered doses requires the vaccination of all individuals aged 50 years or more and the vaccination of at least 30% of younger individuals. In remote settlements, the same achievement would require the vaccination of at least 90% individuals over 50 years of age and a vaccination coverage of 20% in younger ages.

To reduce the cumulative incidence of critical disease under 50 cases per 100,000 individuals in less urbanized areas, a 90% vaccination coverage over 50 years of age should be complemented with more than 70-80% coverage among younger eligible subjects. In urban neighborhoods, the same result would require 90% coverage among the elderly and 50% coverage in younger individuals. If a maximum uptake level of 80% would be achieved in the elderly, to obtain similar results the vaccination of at least 60%, 90%, and 80% of the population under 50 years of age is needed in urban, rural, and remote areas, respectively.

The ranking of different vaccination strategies highlighted under our baseline assumptions is confirmed in a wide spectrum of sensitivity analyses accounting for i) a different efficacy of the vaccine (see Figure S3), ii) the uncertainty in the immunity levels acquired during the first pandemic phase (see Figure S4), and iii) the uncertainty in the reproduction number due to possible changes in the transmission determined by social distancing measures and in the increased transmissibility estimated for Delta compared to pre-circulating lineages (see Figure S5).

## Discussion

A limited vaccine supply should be considered when exploring the impact of vaccination strategies against COVID-19 in low-income countries [1,17]. In this study, we evaluated different age priority targets for vaccination in Ethiopia, considering changes in the disease spread determined by natural immunity acquired during the first year of the pandemic. To this aim, we simulated SARS-COV-2 spread before the launch of the national immunization campaign and assessed the potential disease burden caused by the Delta variant under different vaccination scenarios across urban, rural, and remote areas of the Southwest Shewa Zone.

Obtained results suggest that, before March 2021, infected individuals aged 50 years or more might have been responsible on average for 50.9%, 18.9%, and 15.9% of all critical cases occurred in remote, rural, and urban settings, respectively. Nonetheless, we found that a pivotal role in the spread of SARS-CoV-2 was played by subjects under 30 years, who might have been responsible for about half of the infections in all the considered areas.

Vaccination coverage against COVID-19 has remained extremely low in Ethiopia throughout 2021 [1,17]. As COVID-19 deaths ascertained in this country until December 2021 suggest a mortality rate around 5.9 per 100,000 residents [38], our estimates of the incidence of critical cases in the absence of vaccination highlight that COVID-19 deaths may have been poorly detected in sub-Saharan settings. This is in line with a post-mortem surveillance suggesting 91.4% underreporting of COVID-19 deaths in Zambia [10]. We found that during the Delta epidemic wave less urbanized areas might have been exposed to a higher burden of COVID-19 cases due to older populations or a lower circulation of the infection during the first pandemic year. Additionally, the natural immunity acquired in the first pandemic phase and the reopening of schools significantly increased the proportion of critical cases caused by younger infectors. Nonetheless, our estimates highlight that prioritizing older age segments of the population for vaccination remains the most effective strategy to minimize the burden of critical illness in the Southwest Shewa Zone of Ethiopia. This conclusion emerges irrespectively to the overall number of available doses and despite the high infection rates experienced by the elderly during the first year of the pandemic and the large contribution played by young individuals in the spread of the disease afterwards. Our findings therefore confirm the results obtained across different countries in early 2021 [28,39,40].

Presented results should be interpreted considering the following limitations. The comparison of alternative vaccination priority groups was carried out by assuming that the vaccine is instantaneously administered to all individuals in the target ages, therefore neglecting the time required for the rollout of the vaccination. To better highlight the overall potential of vaccination, we simulated its impact from March 2021, when the national vaccination program was officially launched. Due to the circulation of SARS-CoV-2 after this date and the waning of immunity acquired from natural infection, initial conditions considered to compare the different vaccination strategies do not reflect the current epidemiological conditions in the Southwest Shewa Zone. Nonetheless, the resulting priority ages were found to be robust under alternative modeling assumptions on the immunity level acquired in the first pandemic year and on the vaccine efficacy. It is also worth mentioning that school closure was the only intervention we considered when estimating the age-specific immunity profile before the vaccination launch. This means that variations in the social distancing measures adopted during the first pandemic year were not considered. These include an initial suspension of nonessential productive activities in early 2020 [11] and the progressive re-opening of schools from November 2020 [21,41]. However, the carried-out analysis shows that our model was sufficiently robust to reproduce the age distribution of SARS-CoV-2 infections identified in the considered region during the first wave of COVID-19. As transmission after March 2021 was simulated under the hypothetical scenario of an unmitigated COVID-19 epidemic, our estimates of the expected number of infections and critical cases under different vaccination programs should be considered as illustrative worst-case scenarios to compare the performance of alternative vaccination strategies. Finally, because of the lack of direct data from Africa, the relative susceptibility, the age-specific risks of developing a critical disease, and the potential increased transmissibility and immune escape associated with the Delta variant were assumed from evidence gathered in other countries [4,18,26].

## Conclusions

Despite infections among children and young adults likely caused 70% of critical cases during the Delta wave in SWSZ, most vulnerable ages should remain a key priority target for vaccination against COVID-19. Considering the potential emergence of novel variants of SARS-CoV-2 in the future, our estimates suggest that in Ethiopia older individuals residing in less urbanized settlements should be prioritized for vaccination. Future non-pharmaceutical interventions should focus on reducing potential infectious interactions between the elderly and individuals under 30 years of age, representing their most likely infectors.

## Data Availability

All data produced in the present work are contained in the manuscript

## Declarations

### Ethics approval and consent to participate

Not applicable

### Consent for publication

Not applicable

### Availability of data and materials

Data analysed during this study can be found in [11, 23]

### Authors’ contributions

P.P., M.A., and S.M. conceived the study. A.Z. and M.G. wrote the code and performed the analysis. A.Z., M.G., and P.P. wrote the first draft of the manuscript. P.P., M.A., and S.M. supervised the study. All authors contributed to interpret the results, read, reviewed, and approved the final version and the submission of the manuscript. The corresponding author had final responsibility for the decision to submit for publication.

## Acknowledgements

Not applicable

## Supplementary material

**Figure S1.**
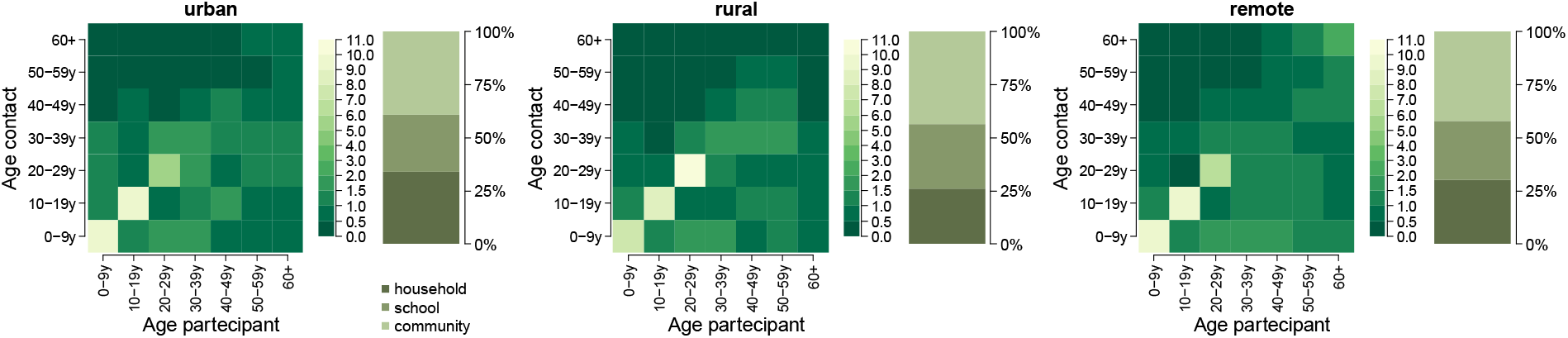
Contact matrices representing the mean number of daily contacts reported by a participant in the age group *i* with individuals in the age group *j* in each site (urban, rural, and remote). The bar plots show the percentage of contacts that occurred in each setting (household, school, and community).

**Figure S2.**
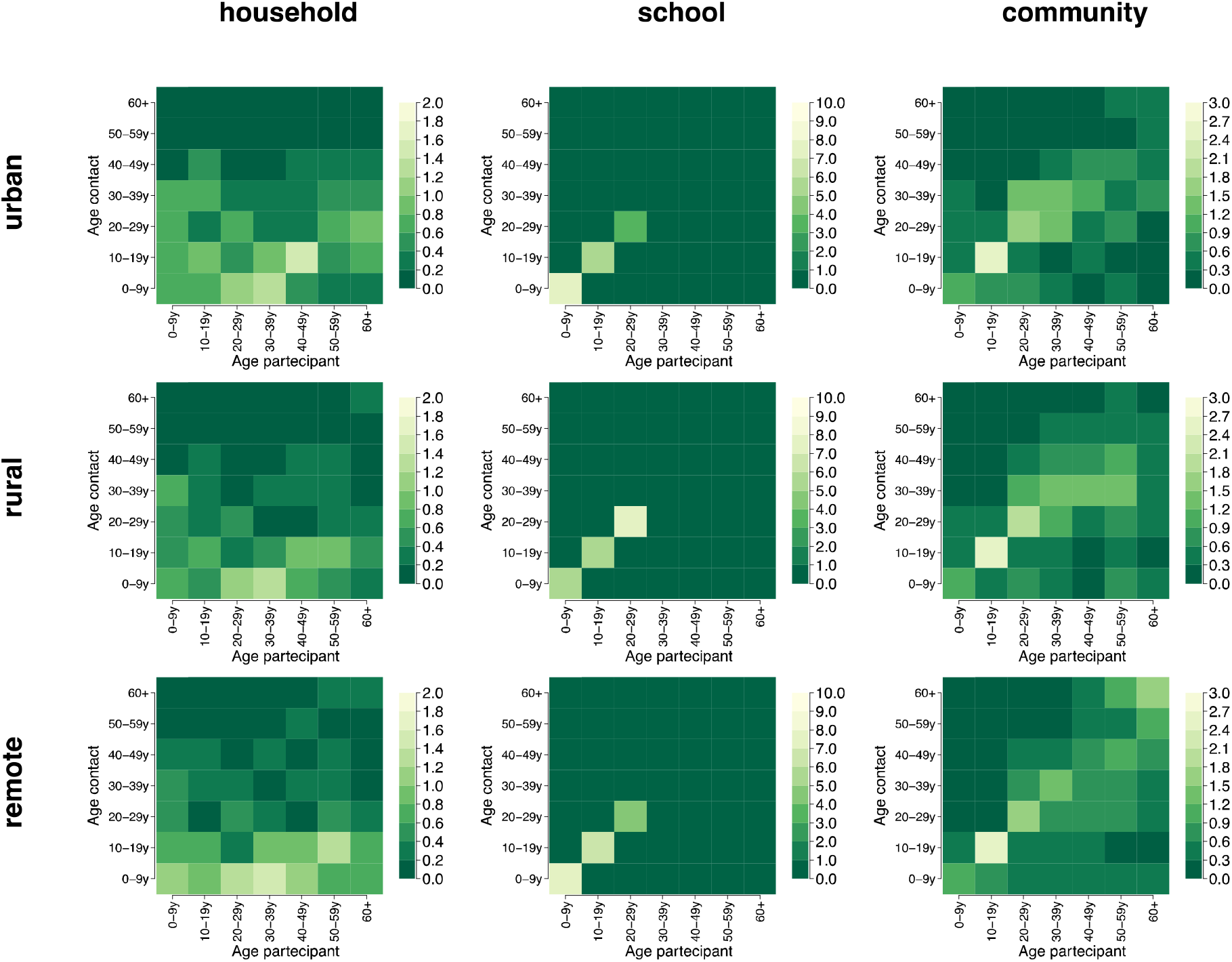
Contact matrices representing the mean number of daily contacts reported by a participant in the age group *i* with individuals in the age group *j* in each setting (household, school, and community) and site (urban, rural, and remote).

**Figure S3.**
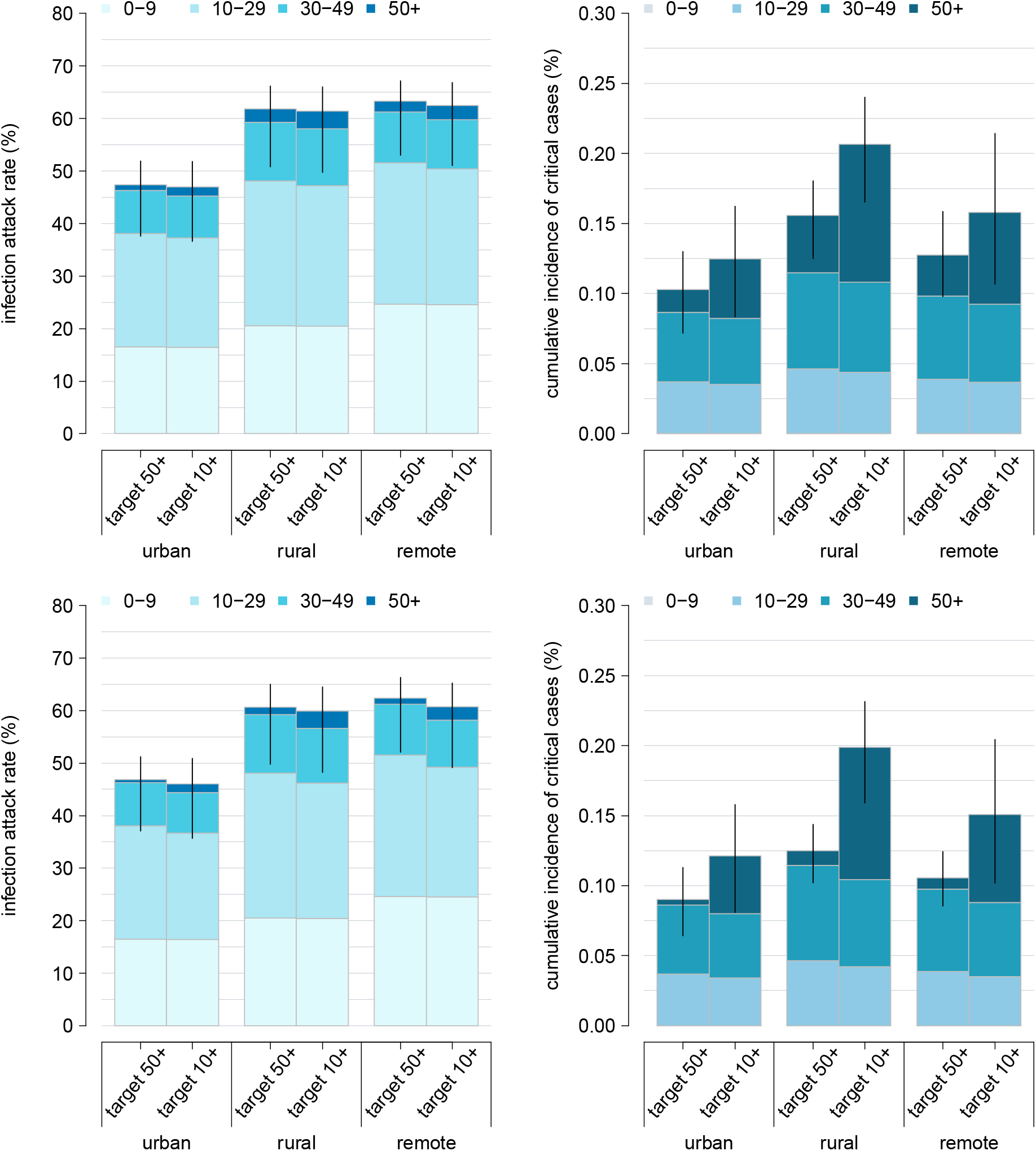
Sensitivity analysis on vaccine efficacy. Estimated infection attack rate and cumulative incidence of critical cases expected across different geographical contexts (urban, rural, and remote), as obtained under the assumption that either all the individuals aged 50 years or older are vaccinated or the corresponding number of vaccine doses is uniformly distributed throughout the population over 10 years. Estimates are obtained assuming a lower vaccine efficacy (set at 55% against infection and 45% against critical disease; first row) and a more effective vaccine (with an efficacy of 80% against infection and 75% against critical disease; second row). Colored bars represent average estimates, stratified by the age group of infected individuals (0-9, 10-29, 30-49, 50+ years); solid lines represent the 95% CI of model estimates.

**Figure S4.**
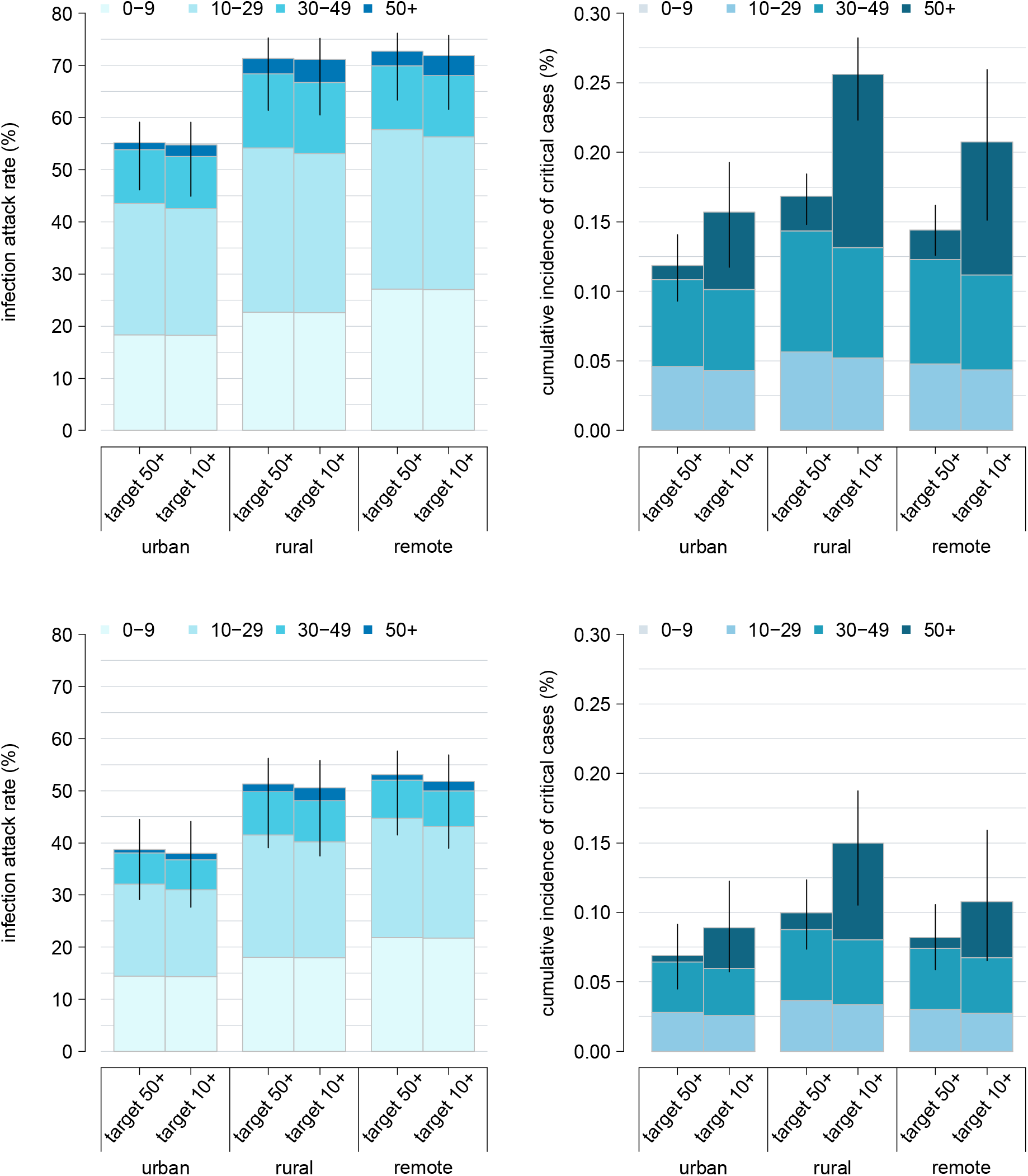
Sensitivity analysis on initial natural immunity. Estimated infection attack rates and cumulative incidence of critical cases expected across different geographical contexts (urban, rural, and remote), as obtained under the assumption that either all the individuals aged 50 years or older are vaccinated or the corresponding number of vaccine doses is uniformly distributed throughout the population over 10 years. Estimates are obtained assuming a lower initial natural immunity (22% in rural and in remote, 38% in urban; first row) and higher initial immunity levels (40% in rural and in remote, 53% in urban; second row). Colored bars represent average estimates, stratified by the age group of infected individuals (0-9, 10-29, 30-49, 50+ years); solid lines represent the 95% CI of model estimates.

**Figure S5.**
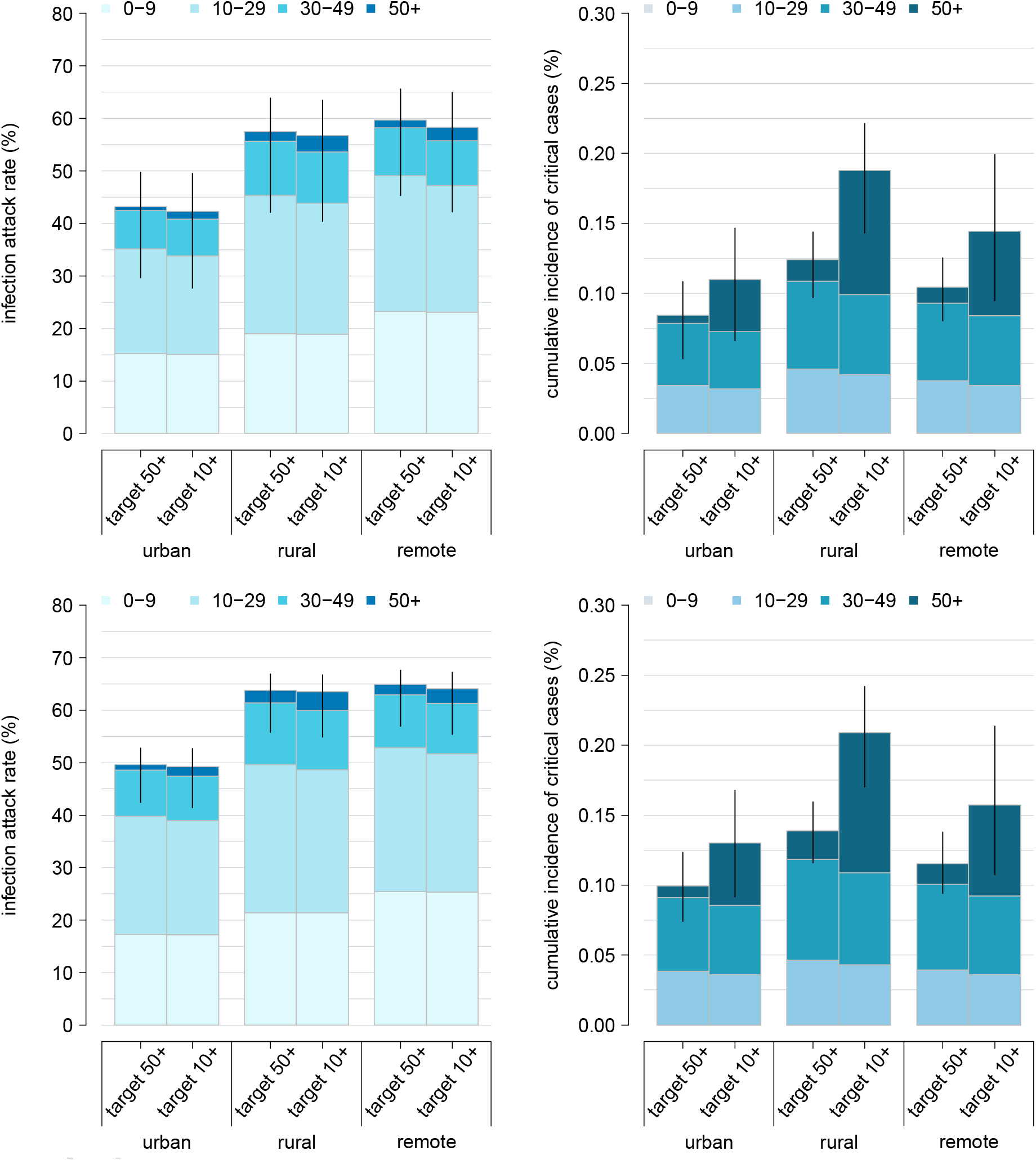
Sensitivity analysis on transmissibility. Estimated infection attack rates and cumulative incidence of critical cases expected across different geographical contexts (urban, rural, and remote), as obtained under the assumption that either all the individuals aged 50 years or older are vaccinated or the corresponding number of vaccine doses is uniformly distributed throughout the population over 10 years. Estimates are obtained assuming a 15% decrease (first row) and a 15% increase (second row) in the SARS-CoV-2 transmissibility. Colored bars represent average estimates, stratified by the age group of infected individuals (0-9, 10-29, 30-49, 50+ years); solid lines represent the 95% CI of model estimates.

